# Impact of non-pharmaceutical interventions targeted at the COVID-19 pandemic on influenza activity in the UK Armed Forces

**DOI:** 10.1101/2022.06.12.22276290

**Authors:** George Otieno, Ngwa Niba Rawlings

## Abstract

**Introduction:** Non-pharmaceutical interventions (NPIs) such as lockdown, social distancing and use of face coverings was adopted by the United Kingdom (UK) Armed Forces (AF) during the COVID-19 pandemic. This study assessed the impact of the use of NPIs on influenza activity in the UK AF.

**Methods:** A longitudinal study design was adopted, and secondary data was analysed retrospectively. Clinical Read codes for influenza-like illness (ILI) was used to generate data for flu seasons before and during the COVID-19 pandemic (September 2017 to April 2021).

**Results:** Before the COVID-19 pandemic, the rate of reporting ILI was ∼ 4% across all flu seasons. The count of ILI was 2.9%, 2.2% and 3.1% during 2017-18, 2018-19 and 2019-20 flu seasons respectively. During the COVID-19 pandemic, both the rate of reporting ILI (0.6%) and the count of ILI (0.5%) were significantly smaller (p < .001). The rate of reporting ILI was positively correlated with the count of ILI (r (2) = .97, p = .014). Influenza vaccination rate increased by 1.3% during the COVID-19 pandemic. vaccination rate was negatively correlated with the rate of reporting ILI (r (2) = -.52, p = 0.24) and the count of ILI (r (2) = -.61, p = 0.19). However, this correlation was not significant. The use of NPIs was negatively correlated with the rate of reporting ILI (r (2) = -.99, p = < .001) and the count of ILI (r (2) = -.95, p = 0.026). The overall multiple regression performed was statistically significant (R^2^ = 0.94, F (1, 2) = 33.628, p = 0.028). The rate of reporting ILI significantly predicted the count of ILI (β = 0.609, p = 0.028) while vaccination rate did not significantly predict the count of ILI (β = -0.136, p = 0.677).

**Conclusions:** Influenza activity in the UK AF was significantly reduced during the COVID-19 pandemic. The use of NPIs and the rate of reporting ILI significantly reduced the count of ILI. Being vaccinated for influenza did not significantly reduce the count of ILI.

## Introduction

Influenza (flu) is an acute respiratory infection caused by the influenza virus, with severe implications among high-risk groups such as children, the elderly, pregnant women, health workers and those with serious medical conditions [1,2]. Like COVID-19 [3,4], influenza is predominantly transmitted from person to person through respiratory droplets when people talk, sneeze or cough [2,5,6]; however, the risk of transmission is increased in places where people stay near each other for prolonged periods such as schools, hospitals, and military barracks. It is estimated that there are over 1 billion cases of influenza annually, with an estimated 3 - 5 million severe cases and 290 - 650 thousand influenza-related respiratory deaths [7]. According to the Office of National Statistics, the number of deaths from influenza in England and Wales was 510 in 2020, compared to 1,213 and 1,596 in 2019 and 2018 respectively [8].

ILI has been defined by the European Centre for Disease Control (ECDC) as a sudden onset of symptoms with at least one of fever (chills); malaise; headache; muscle pain and at least one of cough; sore throat; shortness of breath. Influenza-like illness (ILI) represents a group of symptoms which are similar to those of influenza and caused by more than 200 viruses [9]. Without laboratory confirmation, it is difficult to distinguish between true influenza and ILI [10].

In the military, influenza virus is one of the few infections able to stop military operations due to its ability to cause a large population of healthy soldiers to suddenly become ill. During the 1918-19 influenza pandemic, approximately 50,000 British soldiers were hospitalized in a single week, with an estimated 10,000 deaths [11]. The United Kingdom (UK) Armed Forces (AF) sees two types of influenza commonly reported amongst its personnel during flu season; which are influenza A and influenza B [12]. Rapid Point of Care testing (POCT) based on nucleic acid amplification technologies (NAAT) is commonly used in UK healthcare facilities and in some military medical facilities to test for influenza [12]. Antigen-based rapid influenza detection tests, based on immunological detection of viral antigens are also used. However, laboratory confirmation is not always conducted in medical treatment facilities (MTFs) in the UK AF and influenza is recorded as either total instance of ILI (the rate of reporting ILI recorded over the phone or in person based on syndromic diagnosis) and the count of ILI (number of service personnel who reported ILI).

The UK AF Defence Medical Services offers a yearly Seasonal Influenza immunisation programme which offers vaccinations to eligible military personnel (adults aged between 50-64, healthcare staff who are in direct contact with patients and personnel with underlying health conditions) [13]. As the UK AF population is predominantly made up of fit and younger individuals who are under 50 years of age (healthy adults), those eligible for the influenza vaccination programme make up a very small number. A study by Dermont and Elmer has questioned the justification for offering and expanding the vaccination programme in the UK AF [14]. They augured that the very small proportion of service personnel (SP) who go on to experience influenza are likely to recover uneventfully after a few days of illness. Although the main consequence of influenza in adults is time off work, a 2017 Cochrane review showed that vaccines may lead to little or no small reduction in days off work [15].

COVID-19 and influenza are both respiratory diseases with similar transmission pathways [16]. In the course of the COVID-19 pandemic, influenza activity has remained low as confirmed by multiple national surveillance systems in various countries [17–25]. Public health interventions including non-pharmaceutical interventions (NPIs) targeted at the COVID-19 pandemic have been reported to have impacted influenza activity [17,18,20,26,27]. These NPIs include individual measures (such as hand hygiene, respiratory hygiene and use of face masks); population-related measures (such as promoting physical distancing and restricting movement/ gathering of people); and environmental measures (such as cleaning and ventilation of indoor spaces) [28].

NPIs used in the UK to control COVID-19 include public health messaging, social and physical distancing measures, national lockdowns (with most people working from home), the wearing of face coverings, hand hygiene and travel restrictions [17]. These measures were adopted in military facilities, including other NPIs such as capacity restrictions in offices and communal areas, use of screens to separate workstations in offices, isolation of cases and contacts, use of posters to educate and remind soldiers about COVID-19 prevention measures, conducting military training in smaller cohorts/teams and regular cleaning/disinfecting surfaces and high touch points such as door handles and communal items. Although these measures aim to control COVID-19 transmission, they potentially had similar effects on influenza activity. This study aimed to assess the impact of the use of NPIs targeted at the COVID-19 pandemic on influenza activity in the UK AF.

## Materials and methods

### Study population and design

The study population was UK AF (Army, Air Force and Navy) SP either currently serving or who had previously served, with a record of ILI during the study period. A longitudinal study design was used and involved the retrospective analysis of data obtained from the UK AF Defence Medical Information Capability Programme (DMICP).

### Data management

Observational data was obtained from DMICP. Clinical Read codes for ILI using a methodology adapted from Public Health England’s (PHE) in-hours syndromic surveillance as described by Dermont and Elmer [9]. In summary, this methodology (adapted from PHE real-time syndromic surveillance team [29]) included data captured from Read-coded entries in DMICP to assess the evidence for ILI in UK AF SP for the period from September 2017 to April 2021. Multiple searches were conducted for all unique patient identifiers with accompanying Read codes associated with ILI. Duplicate entries were removed and only the first occurrence of ILI was included.

Data on ILI used for this study covered flu seasons (September to April) from 2017 to 2021. The data included the count of ILI before the COVID-19 pandemic (September 2017 – April 2018, September 2018 – April 2019; September 2019 - April 2020) and during the COVID-19 pandemic (September 2020 – April 2021). Sensitive information such as name, gender, age, and location were not included in the data provided. Data on influenza vaccination rate and the rate of reporting ILI during the study period in the UK AF was also collated and analysed. The data was checked for validity and entered into Microsoft Excel to facilitate storage and analysis. The data was uploaded onto IBM SPSS version 27 and analysed. Data analysis involved both descriptive and inferential statistical computations. Descriptive and inferential statistics and p-value of < 0.05 was considered statistically significant.

### Ethical considerations

This study was guided by the Helsinki Declaration as revised in 2013. Ethical approval was obtained from Leeds Beckett University ethics committee in line with its ethics policy and procedures. Authorisation was also obtained from the UK AF who provided the secondary data used for this study. Consent was waved as no sensitive information was included in the data.

## Results and discussion

### ILI in the UK AF

Before the COVID-19 pandemic, the rate of reporting ILI was ∼ 4% across all three flu seasons, occurring in 4254 (2.9%), 3258 (2.2%) and 4508 (3.1%) of SP in 2017-18, 2018-19 and 2019-20 flu seasons respectively. During the COVID-19 pandemic (2020-21 flu season), the rate of reporting ILI was 0.6%, occurring in 791 (0.5%) SP (Fig 1). There was homogeneity of variances, as assessed by Levene’s test for equality of variances, for competence, p > .05. A two-sample t-test was performed to compare the count of ILI in flu seasons before and during the COVID-19 pandemic. There was a significant difference in the count of ILI between flu seasons before the COVID-19 pandemic; mean (M) = 494.25, SD = 300.66 and during the COVID-19 pandemic (M = 95.38, SD = 70.56); t (14) = 3.63, p = < 0.001.

**Fig 1.**
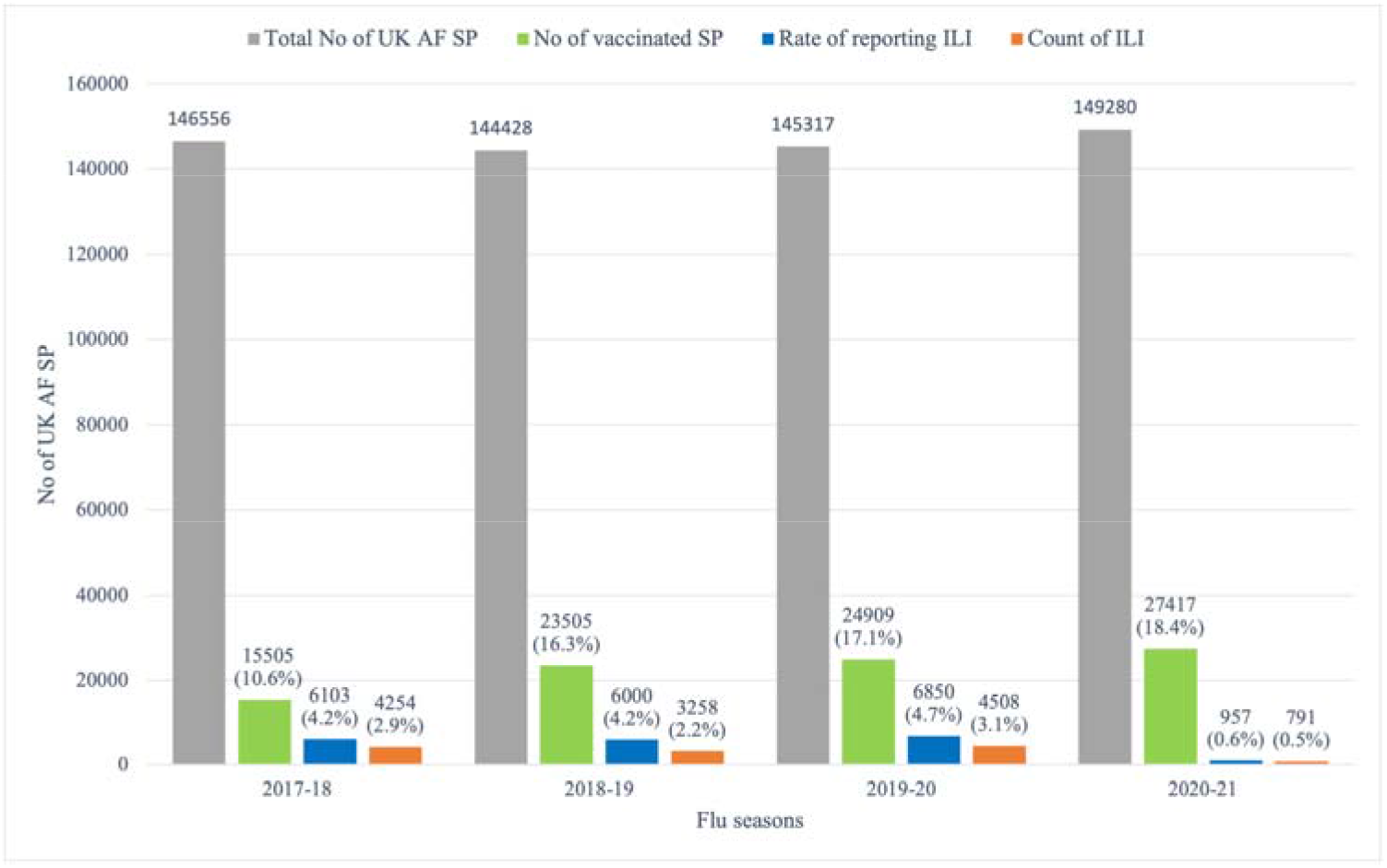
Total number of SP; vaccination rate; rate of reporting ILI; and count of ILI in the UK AF before and during the COVID-19 pandemic.

The results showed a significant reduction in influenza activity during the COVID-19 pandemic in the UK AF. Similar findings have been reported by surveillance systems in the UK [17], United States [25], New Zealand [18], Australia [24], Korea [22,23], Taiwan [21] and China [19,20,27]; which have all shown a significant reduction in influenza activity (transmission, number of cases and burden of disease) during the COVID-19 pandemic. These reports attribute the reduction in influenza activity to the implementation of various NPIs. The use and stringent adherence to NPIs during the COVID-19 pandemic in the UK AF potentially reduced influenza activity. The count of ILI before and during the COVID-19 pandemic showed a positive correlation, with a steady reduction from January to April every flu season except for the 2019-20 season. During the 2019-20 season (onset of the COVID-19 pandemic), there was a rise in the count of ILI from February to March 2020 and a steep decline in the count of ILI (from 781 to 29 SP) from March to April 2020 (Fig 2). This inconsistency in the data trend occurred during the period of the first national lockdown in the UK [30]. A study conducted in Canada had similar findings, showing a drastic decrease in influenza A and B laboratory detections concurrent with social distancing measures and nonpharmaceutical interventions [31]. Another study conducted in the United States also had similar findings as influenza virus circulation declined steeply within 2 weeks of the COVID-19 emergency declaration and widespread implementation of community mitigation measures [32].

**Fig 2.**
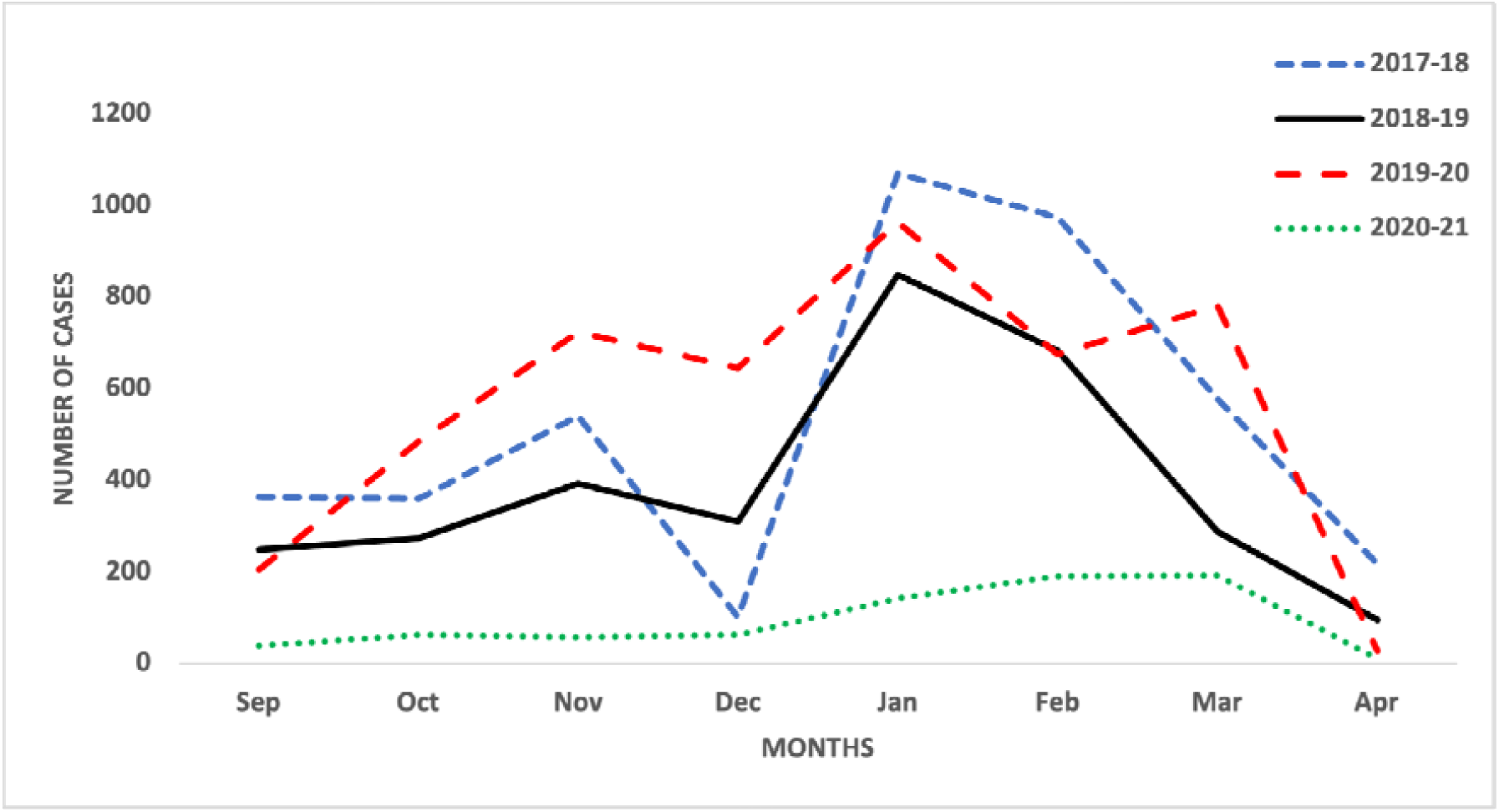
Monthly count of ILI in the UK AF before and during the COVID-19 pandemic.

The rate of reporting ILI in the UK AF was significantly lower during the COVID-19 pandemic (Fig 1). A Pearson correlation coefficient was computed to assess the linear relationship between the rate of reporting ILI and the count of ILI. There was a positive correlation between the two variables, r (2) = .97, p = .014 (Fig 3). This meant that a decrease in the rate of reporting ILI resulted in a corresponding decrease in the count of ILI and vis versa. During the implementation of a national lockdown, majority of UK AF SP were working from home; person to person contact was reduced, preventing the spread of the influenza virus. This led to a significant reduction in the rate of reporting ILI and subsequently a reduction in the count of ILI. Even though many studies have attributed the decrease in influenza activity during the COVID-19 pandemic to the implementation of NPIs [17–25], a study conducted in the US linked the reduction in rate of influenza during the 2019-2020 influenza season to change in public behaviour [33]. However, the study agreed that this change in behaviour was because of the implementation of public health interventions (NPIs).

**Fig 3.**
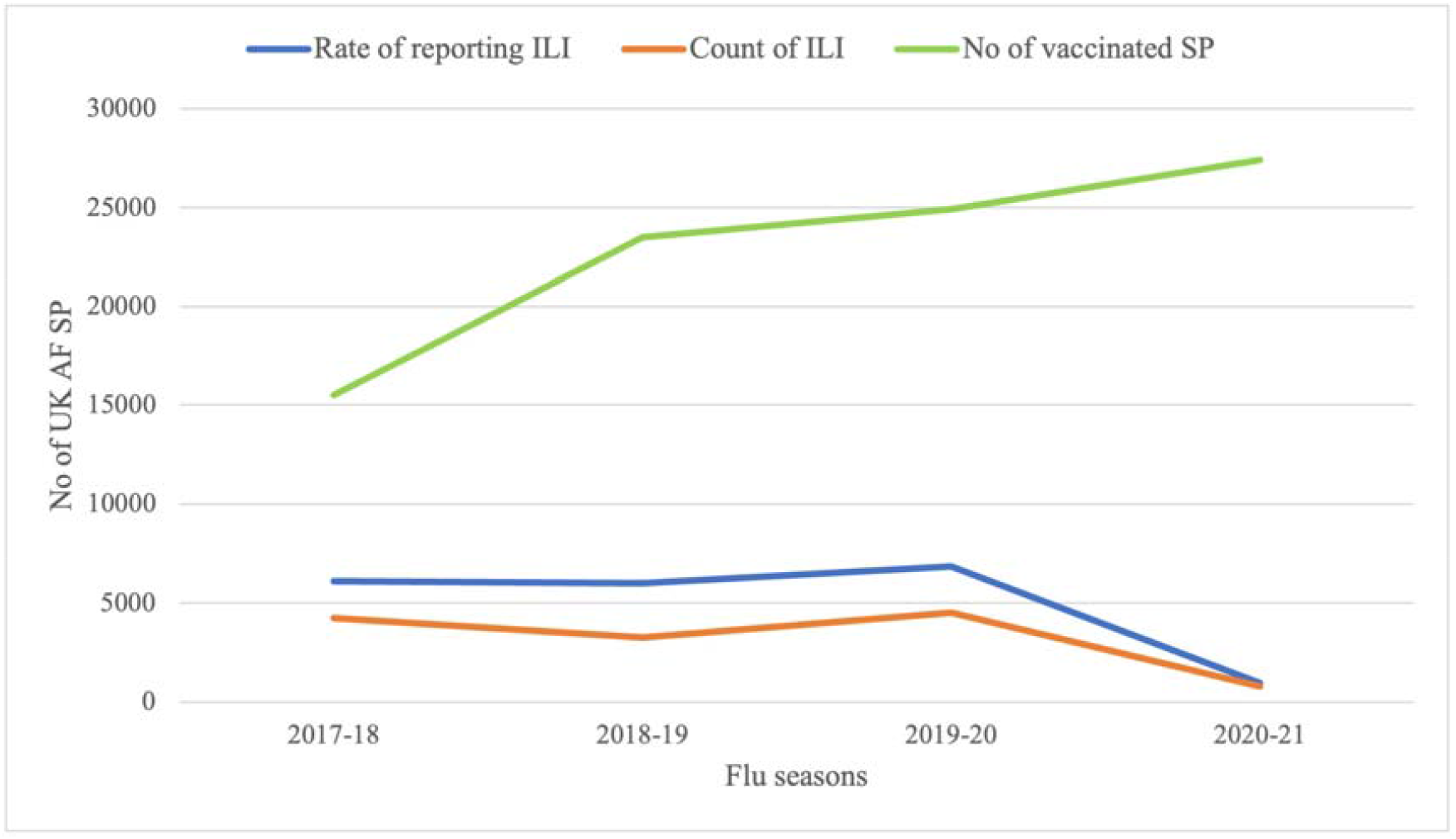
Relationship between the rate of reporting ILI, the count of ILI and vaccination rate in the UK AF before and during the COVID-19 pandemic.

The use of NPIs was coded to determine its relationship with the rate of reporting ILI and the count of ILI. A Pearson correlation coefficient was computed to assess the linear relationship between {i} use of NPIs and the rate of reporting ILI and {ii} use of NPIs and the count of ILI. There was a negative correlation in both instance between the two variables, {i} r (2) = -.99, p = < .001 and {ii} r (2) = -.95, p = 0.026. Therefore, the use of NPIs significantly reduced influenza activity (both the rate of reporting ILI and the count of ILI). According to the UK Health Security Agency’s syndromic surveillance, NPIs implemented during the COVID-19 pandemic reduced the transmission of a range of respiratory pathogens resulting in ILI (including flu, rhinovirus, and other Corona viruses) [29]. Although the implementation of NPIs significantly reduced influenza activity, other factors related to the management of COVID-19 in the UK AF potential impacted influenza activity as well. For instance, during the COVID-19 pandemic, a majority of SP were working from home due to the national lockdown. They were less likely to report flu-like symptoms to MTFs and only got tested for COVID-19 generally because they felt it was not relevant or they didn’t wish to be tested and required to self-isolate. Moreover, MTFs being burdened with COVID-19 cases did not routinely screen for ILI as they would normally do on SP with flu-like symptoms before the pandemic. COVID-19 reporting in the UK AF was done using mechanisms such as the COVID-19 Reporting Tool, MyRAF and MyNavy which mandated weekly health reporting and may have meant that SP did not border to report their symptoms to MTF after using these tools. The fear of testing positive for COVID-19 might have caused some SP not to report their flu-like symptoms to MTFs especially because influenza and COVID-19 are symptomatically indistinguishable. However, a study conducted in south-eastern Wisconsin looking at decrease in positivity rate of influenza tests coinciding with outbreak of SARS-CoV-2 concluded that it is highly unlikely that patients with influenza decided to avoid seeking medical care [33].

### Influenza vaccination in the UK AF

The number of UK AF SP who were vaccinated during the COVID-19 pandemic was higher than those vaccinated before the COVID-19 pandemic by 1.3%, which was not substantially different across flu seasons (Fig 1). A Pearson correlation coefficient was computed to assess the linear relationship between {i} vaccination rate and the rate of reporting ILI and {ii} vaccination rate and the count of ILI. There was a negative correlation in both instance between the two variables, {i} r (2) = -.52, p = 0.24 and {ii} r (2) = -.61, p = 0.19 (Fig 3). These results showed that an increase in the vaccination rate should cause a corresponding decrease in the rate of reporting ILI and the count of ILI. However, the relationship not being significant means there is a probability of this occurring by chance. Therefore, increasing the vaccination rate for influenza in the UK AF SP will not necessarily cause a reduction in influenza activity (the rate of reporting ILI and the count of ILI).

These findings were consistent with those of Flannery and colleagues who suggested that vaccination did not significantly reduce medically attended influenza illness due to A(H3N2) virus infection [34]. To some extent, our findings also agree with the 2017 Cochrane review which stated that ‘…. vaccines may lead to little or no small reduction in days off work’ [15], and with Dermont and Elmer who questioned the impact and relevance of influenza vaccination on a fit population such as the UK AF in keeping SP at work or returning them to work any sooner [14]. In comparing which one of two interventions (vaccination and use of NPIs) would reduce influenza activity, we can to some extent rule out vaccination as an effective intervention in otherwise fit individuals such as in the UK AF. Therefore, the use and stringent adherence to NPIs would be a preferable intervention in reducing influenza activity. However, a combination of both interventions could have an even greater effect in reducing influenza activity.

Three independent (predictor) variables (use of NPIs, the rate of reporting ILI and vaccination rate) were initially included in the linear regression analysis to understand how much effect they had on the dependent (outcome) variable (the count of ILI). Assumption testing of the predictor variables showed multicollinearity amongst two of the variables (use of NPIs and rates of reporting ILI) as the correlation coefficient (r) between the two predictor variables was > 0.7. This meant that both predictor variables would have a similar effect on the outcome variable, and therefore, one of them had to be removed from the regression analysis. The multiple linear regression performed was used to test if the rate of reporting ILI and vaccination rate significantly predicted the count of ILI. The fitted regression model was: the count of ILI = 173.13 + 0.609* (the rate of reporting ILI) – 0.136* (vaccination rate). The overall regression was statistically significant (R^2^ = 0.94, F (1, 2) = 33.628, p = 0.028). It was found that the rate of reporting ILI significantly predicted the count of ILI (β = 0.609, p = 0.028); while vaccination rate did not significantly predict the count of ILI (β = -0.136, p = 0.677).

According to the above multiple regression analysis, the rate of reporting ILI was a significant predictor of the count of ILI; meaning that the use of NPIs would also significantly predict the count of ILI because both predictor variables expressed collinearity based on assumption testing. This analysis confirms the initial findings showing that out of the three independent variables, the rate of reporting ILI and the use of NPIs had a significant effect on the count of ILI while vaccination rate will not.

### Limitations

These findings cannot be generalized to pandemic influenza given that our population (military personnel) are generally fit, unlike a normal community population and the underlying population immunity to seasonal influenza. Laboratory tests are not always used to confirm ILI in the UK AF. A majority of the ILI recorded is based on syndromic diagnosis and as a result, some proportion of the count of ILI recorded will not be true influenza. As it was not possible to exclude that proportion from the analysis, it is understood that it would have some influence on the results. This study did not measure how much each independent (predictor) variable affected the dependent (outcome) variable. This would be a viable area requiring further research. Also, further studies could look at the impact of individual NPIs and assess suitable NPI combinations with the most effect on reducing influenza activity.

## Conclusions

Analysis of DMICP data on ILI in the UK AF showed a significant reduction in influenza activity during the COVID-19 pandemic. The use of NPIs significantly contributed to the reduction in the count of ILI. This findings has been corroborated by various studies demonstrating that NPIs significantly reduce influenza activity (transmission, number of cases and burden of disease) during the COVID-19 pandemic [17–25]. However, in the UK AF, the rate of reporting ILI (total instances of ILI) was also a significant predictor of the count of ILI. Although vaccination rate was correlated with the count of ILI, this relationship was not significant. Being vaccinated for influenza in the UK AF did not significantly reduce influenza activity (the rate of reporting ILI and the count of ILI). Nevertheless, influenza vaccination has been shown to reduce severe flu illness, hospitalisation, and death amongst people with a higher risk of flu complications such as pregnant people, young children, adults 65 years and older and adults with certain chronic health conditions [35].

## Data Availability

All relevant data are within the manuscript and its Supporting Information files.

## Acknowledgements

We would like to acknowledge the Defence Public Health Unit, the Defence College of Health Education/Training, the Defence Environmental Health Cadre and Leeds Beckett University for facilitating this study.

## Supporting information

**S1 Fig.**
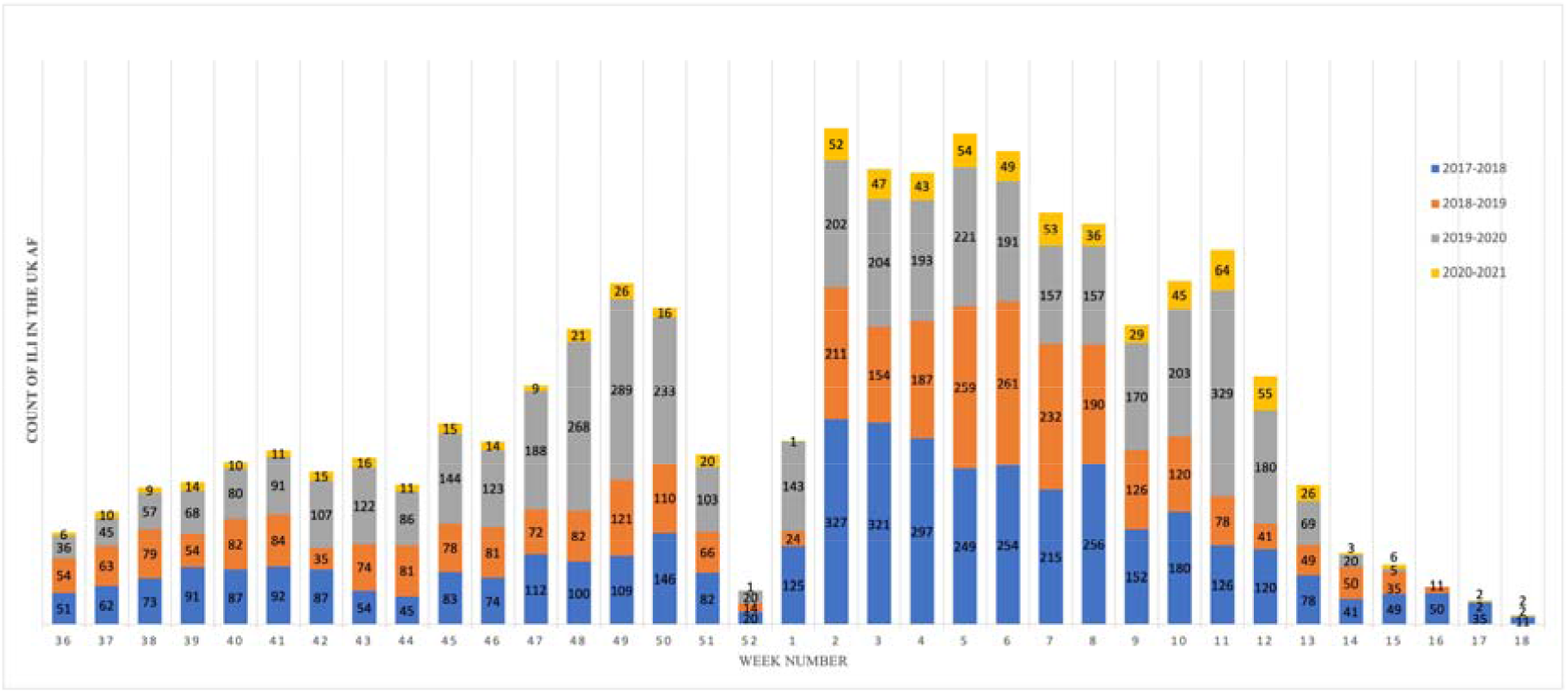
Weekly count of ILI in the UK AF (2017 – 2021 flu seasons)

